# Validity, Reliability, and Framing Effects in Equity-Efficiency Trade-Off Studies: A Systematic Review

**DOI:** 10.1101/2024.10.14.24315354

**Authors:** Victoria Chechulina, Andrew Chu, Quang Hung Lam, Camryn Kabir-Bahk, Shehzad Ali

## Abstract

**Objectives:** Health system resources are limited, and decision makers often need to make trade-offs between equity and efficiency. Such trade-offs are guided by public values that are elicited using choice experiments. There is no standard approach to elicit equity-efficiency trade-offs. Previous studies have found significant variability in public values, raising concerns about the robustness of trade-off experiments. The objective of this systematic review was to determine whether and how equity-efficiency trade-off studies consider validity, reliability and framing effects.

**Methods:** We searched Medline, EMBASE, and Web of Science in January 2024 for health-related equity-efficiency trade-off studies. Two reviewers independently screened titles and abstracts, followed by full-text review, data extraction, and quality assessment.

**Results:** 115 equity-efficiency trade-off studies were identified, of which 29 studies (25.2%) investigated validity, reliability, and/or framing effects. 14 (12.2%) studies assessed validity, 9 (7.8%) studies assessed reliability, and 10 (8.7%) assessed framing or cognitive effects. Validity was most frequently assessed by comparing results to hypothesized expectations, while reliability was commonly assessed by providing a repeated test or questionnaire. Framing and cognitive effects were assessed by varying question order or changing the wording or framing of the scenario. 19 of 23 studies reported high or acceptable validity or reliability, and 5 of 10 studies found no significant framing or cognitive effects.

**Conclusion:** This systematic review highlights the need to consider robustness of elicited values in choice experiments. It will also guide future choice experiments that aim to estimate inequity aversion.

## Introduction

Health equity is a major public policy concern; however, limited progress has been made in recent decades.^1,2^ Health inequities can be mitigated through fairer allocation of health system resources.^3,4^ The standard economic evaluation framework used for resource allocation decisions is based on the efficiency principle which asserts that decisions should maximize the overall health, irrespective of its distribution.^5^ In contrast, the equity principle focuses on fairness in the population distribution of health.^6^ Lately, alternative economic evaluation approaches have been proposed, such as the distributional cost-effectiveness analysis (DCEA), that explicitly incorporate equity concerns in decisions.^7,8^ These approaches use evidence on public values concerning the trade-offs between health maximization (efficiency) and health equity to inform the optimal allocation of resources. Incorporating public values allows decision-makers to explicitly and transparently consider equity in decision-making.

To elicit public preferences on equity-efficiency trade-offs, preference elicitation studies are frequently used in the health economics literature.^9^ These studies typically present individuals with hypothetical scenarios where they are asked to choose between different levels of health gains and their distribution across social groups, in turn revealing the level of inequity aversion of the respondent.^10^ Previous systematic reviews of equity-efficiency trade-off studies have summarized public values on inequity aversion.^11–13^

There are no standard questionnaire instruments to elicit equity-efficiency trade-offs. Studies in the literature have used different questionnaire designs and have estimated different levels of inequity aversion.^11–13^ For example, in a systematic review of equity-efficiency trade-off studies based in the UK, McNamara et al (2020) identified eight studies reporting evidence of inequality aversion, two finding no aversion, and five reporting mixed evidence.^11^ The estimated Atkinson inequality aversion parameter ranged between 1 (i.e. no aversion) and 28.9, and the weight placed on a marginal gain to group with low lifetime health ranged between 1 and 166.2. While this large difference in inequity aversion in studies conducted in the same country can partly be explained by between-study heterogeneity, this degree of variation poses a challenge in determining which estimate can be reliably used for policy decisions.

One consideration that has received limited attention in the literature is the validity, reliability, and framing effects in equity-efficiency trade-off studies. While validity and reliability are frequently studied in relation to questionnaire instruments, such as quality-of-life tools; however, they are less frequently reported for questionnaire-based choice experiments.^17^ While the trend is changing with a number of discrete choice experiments explicitly investigating reliability, validity and framing effects,^18–20^ their use in equity-efficiency trade-off experiments has not been studied.

Janssen et al (2017) have developed a conceptual framework to study validity and reliability of choice experiments, focusing on the measurement and choice-related domains.^21^

To investigate validity, it is important to consider the normative, value-based nature of equity-efficiency trade-off experiments which is distinct from conventional discrete choice experiments where the expected direction of attribute preference can often be reasonably assumed (e.g. more health benefit being better than fewer). While not all aspects of validity are pertinent to equity-efficiency trade-off experiments, several domains may be relevant, including face validity, convergent validity, external validity of measurement, monotonicity, and non-compensatory choices. Face validity of responses can be assessed by examining deviations from hypothesized response behaviors,^22^ such as the level of pro-rich or extreme egalitarian values among respondents,^14^ the willingness-to-pay for larger-sized quality of life gains, or treatment preferences for patients that are younger and have no comorbidities. Convergent validity of the elicited values can be assessed by comparing within or between-subject responses to other choice scenarios/instruments measuring the same underlying construct. For instance, an investigator may compare individual responses against their own social values as measured by social attitude questions within the same survey. Alternatively, the elicited values can be compared with previously published public attitude surveys. Other forms of validity, such as external validity, are more difficult to assess due to limited opportunity to compare elicited values with the predicted or observed choices in the real-world.^21^ Monotonicity relates to consistency in the direction of responses across choice sets. In case of equity-efficiency trade-off studies, this may relate to consistency in relation to a respondent’s preference for welfare increasing choice options.^14^ Finally, non-compensatory choices imply that individuals might only consider one attribute to make choices in the experiment. In case of equity-efficiency trade-off studies, this relates to giving exclusive consideration to the equity attribute such that no level of efficiency loss would compensate for inequity. This may present as an ‘extreme egalitarian’ position when a respondent treats equity as a ‘sacred value’.^14^

Reliability in this context refers to the ability of a measure to produce consistent results over time and space.^23^ While not all domains of reliability are pertinent to equity-efficiency trade-off experiments, test-retest reliability, version consistency and level recoding can be relevant. Test-retest reliability is typically assessed by administering the questionnaire twice, at two different timepoints, assuming stability of preferences.^21^ Version consistency assesses if preferences remain stable across different versions of the questionnaire administered to separate groups of participants.^21^ Finally, level recoding refers to the extent to which participants process the numeric values presented in attributes. To assess this, researchers can administer two versions of the survey with different level values of the health benefits being traded. However, if the units or the range of health benefits are changed across versions, such as changing healthy years to healthy months, the level of trade-offs can be expected to be different.^14^

In addition to validity and reliability, other areas considered in choice experiments are framing and cognitive effects. Framing effects occurs when preferences vary based on how choice scenarios are presented to participants.^14^ The framing may vary in terms of the presentation of scenarios, such as gains versus losses, relative versus absolute changes and small versus large units (such as days versus weeks). For instance, Howard et al (2009) found that framing the accuracy of a cancer detection test in terms of cases found versus cases missed can significantly influence the trade-offs.^24^ Similar framing effects were reported by Veldwijk et al (2016) in relation to the outcome of genetic testing expressed either as survival (positive framing) or mortality (negative framing).^25^ In the context of equity-efficiency trade-off literature, Ali et al (2017) found that the level of inequity aversion varied based on abstract (i.e. not specific to a health condition) versus concrete (i.e. condition-specific) scenario descriptions.^14^ This could be attributed to the idea of agentic responsibility when a specific condition is presented compared to wider structural determinants of health when considering broad health system inequities. Framing effects may violate the principle of invariance, which states that decisions should not vary as a function of equivalent descriptions of the same situation.^26^

Preferences may also vary across scenarios based on the cognitive burden of choice experiments. For instance, probabilistic outcomes may be harder to process cognitively in a choice experiment than fixed frequencies.^27^ As such, the cognitive load of the choice experiment may impact the elicited values.^28^ Researchers should determine whether the results of their preference elicitation studies have been skewed by framing effects or other cognitive biases. Results that have been influenced by cognitive biases may not be reliable enough to inform health policy making, as they may not generalize from the study scenario to real-world settings.^14^

Choice experiments may also employ other approaches to assess the robustness of responses. This may include sensitivity analysis, whereby researchers may exclude participants who did not understand the survey, provided inconsistent responses, or who showed no real preference on the choice task.^29–31^ Such analyses lend credibility to the estimates.

While several systematic reviews of equity-efficiency trade-off studies have been published,^9,11–13,32^ to our knowledge, there has been no systematic review that has examined the validity, reliability or framing effects of equity-efficiency trade-off studies. This review aims to fill this important gap in the literature. If the results of equity-efficiency trade-off studies are to be used in health resource allocation decisions, policymakers need to know if the elicited inequity aversion parameters have been robustly estimated. This study will also provide important methodological insights to improve the design and conduct of future equity-efficiency trade-off studies.

## Methods

This systematic review was designed and reported according to the Preferred Reporting Items for Systematic reviews and Meta-analyses (PRISMA) 2020 guidelines.^33^

### Eligibility Criteria

Study inclusion criteria were determined prior to study screening. We included studies that were: (1) peer-reviewed; (2) elicited value judgements and preferences of individuals with respect to health system allocation decisions or asked individuals to make a health-related equity-efficiency trade-off; (3) included equity or equality as a dimension of choice; and (4) were in English. All stated preference studies were eligible regardless of the elicitation technique. Studies that were theoretical in nature, did not consider health, did not include equity or equality as a dimension of choice, or did not include trade-offs were excluded. There were no restrictions placed on the year of publication, study sample, or study setting. Peer-reviewed original research was included, and conference abstracts were excluded.

### Information Sources

Three databases were searched on December 4^th^, 2023: Ovid MEDLINE (1946 – 04/12/2023), Ovid EMBASE (1947 – 04/12/2023), and Web of Science (1900 – 04/12/2023). Searches were updated on January 12^th^, 2024. Reference lists of systematic reviews of equity-efficiency trade-off studies were manually searched to identify potentially eligible studies.^11–13^

### Search Strategy

The search strategy was designed to capture health-related equity-efficiency trade-off studies by combining three concepts: *health* AND *preference elicitation study* AND *equity or equality*. Terms for each of these three concepts were identified by consulting previous systematic reviews of equity-efficiency trade-off studies.^11–13^ No limits were placed on date or language. The sensitivity of the search strategy was validated by ensuring that studies included in previously published systematic reviews were captured by the search strategy. The full search strategies can be found in Appendix A.

### Study Selection

Records identified from the searches were uploaded to Covidence for screening. Two reviewers (VC, AC, QHL, CKB) independently screened all titles and abstracts, followed by full-text review. Conflicts were resolved by a third reviewer (AC, QHL, CKB). Two reviewers (VC, AC, QHL) independently extracted data from each included study. Discrepancies were discussed with a third reviewer (VC, AC, QHL).

### Data Items and Outcomes

We assessed the proportion of equity-efficiency trade-off studies that evaluated validity, reliability, and/or framing effects. Reviewers then identified how these were assessed, and what the findings were. If a study provided quantitative values for validity, reliability, or framing effects, these values were extracted. If a study discussed them narratively, this information was synthesized.

Data were collected for the following study characteristics in studies that assessed validity, reliability, or framing effects: elicitation method; sample size; geographic location; type of sample (e.g., general public, policymakers, healthcare professionals, etc.); and survey administration method (e.g., face-to-face interview, online survey, etc.).

### Study Quality Assessment

Study quality was assessed using the PREFS checklist, which is suitable for assessing data quality of preference elicitation studies.^34^ The checklist assesses five study domains: purpose; respondent sampling; explanation of methods; findings; and significance testing. Each item is scored 0 (unacceptable) or 1 (acceptable) and each study receives a score from 0 to 5.^35^ Quality assessment was conducted for studies that assessed validity, reliability, and/or framing effects, independently by two reviewers (VC, AC, QHL), and discrepancies were resolved with a third reviewer (VC, AC, QHL). A narrative synthesis was conducted to describe the reporting of validity, reliability, and framing effects in the equity-efficiency trade-off literature.

## Results

The database searches returned 5279 results (1273 MEDLINE, 1424 EMBASE, 2582 Web of Science) and hand searching of reference lists returned three additional citations. After deduplication, 3240 titles and abstracts were screened. The full texts of 200 articles were reviewed, and 115 studies were included. The most common reason for exclusion was the study not being a preference elicitation study (n = 39). The study identification, screening, and inclusion process is summarized in a PRISMA flow diagram (Figure 1).^33^

**Figure 1.**
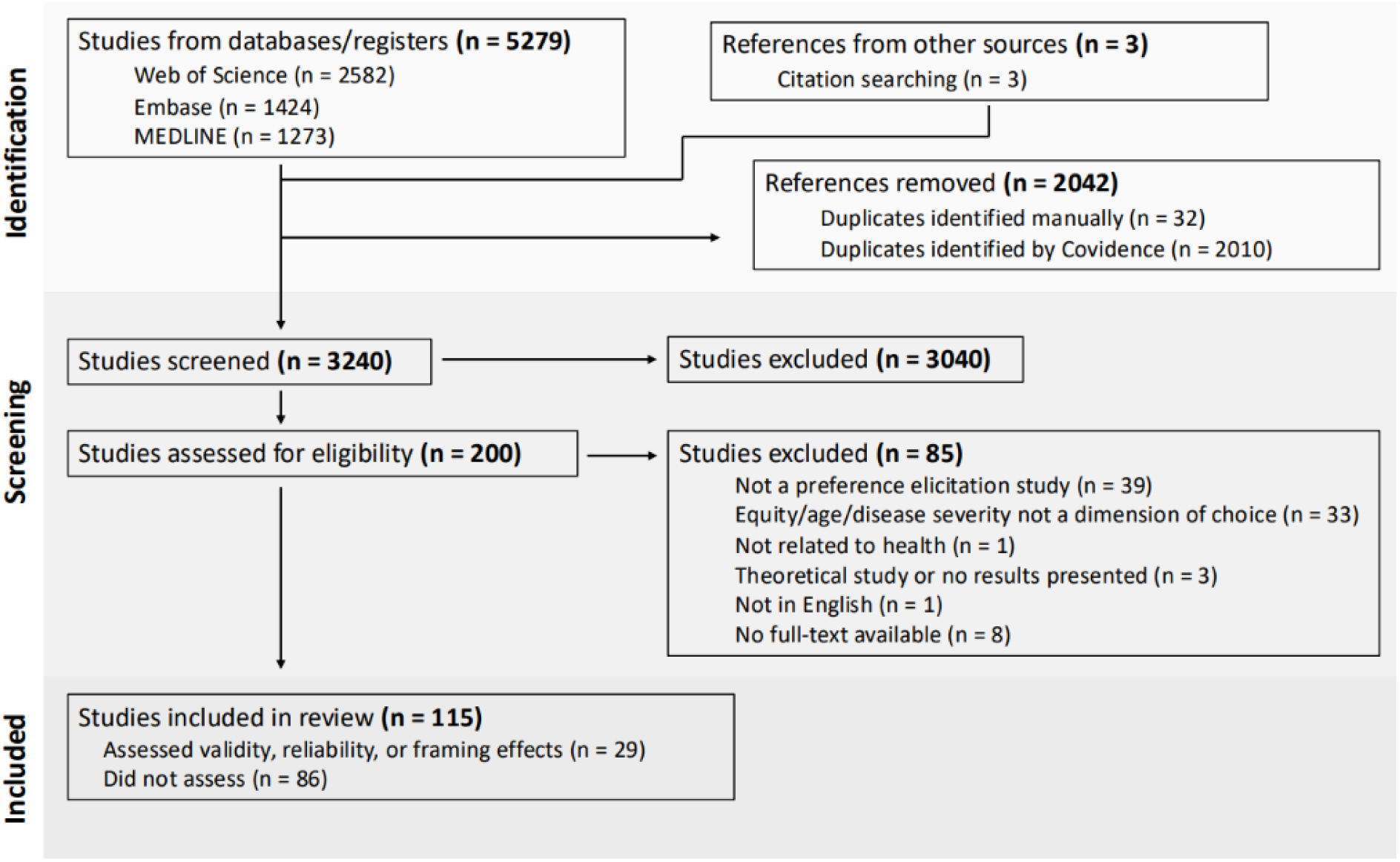
PRISMA flow diagram

Study design characteristics of the studies that assessed validity, reliability, or framing effects are summarized in Table 1. Of the 115 included studies, 29 (25.2%) assessed validity, reliability, or framing effects.^14,26,30,36–61^ The other 86 included studies did not assess or discuss them.^15,16,29,31,62–143^ 14 studies assessed validity (12.2%), 9 studies assessed reliability (7.8%), and 10 assessed framing or cognitive effects (8.7%).

**Table 1.** Study design characteristics of studies that assessed validity, reliability, or framing effects.

| Study | Preference elicitation method | Sample size | Geographic location | Type of sample | Administration format | Property evaluated |
| --- | --- | --- | --- | --- | --- | --- |
| Gulácsi 2012 | Discrete choice experiment | 200 | Hungary | General practitioners | Face-to-face interviews and online questionnaires | Validity |
| Attema 2022 | Person trade-off | 500 | The Netherlands | National sample | Online | Reliability |
| Stolk 2005 | Priority ranking experiment | 65 | The Netherlands | Students, researchers, health policy makers | Not stated | Framing effects and reliability |
| Ratcliffe 2000 | Conjoint analysis | 303 | Britain | Local sample of university employees | Questionnaire sent by mail | Validity |
| Werner 2009 | Survey | 624 | North Israel | Local sample of community-dwelling adults | Face-to-face interview | Reliability |
| Luyten 2019 | Discrete choice experiment | 750 | Belgium | National sample | Survey (not stated whether online or in-person) | Reliability |
| Damschroder 2004 | Person trade-off | 95 | Philadelphia, USA | Local sample (prospective jurors) | Computer-administered survey and face-to-face interview | Reliability and validity |
| Jehu-Appiah 2008 | Discrete choice experiment | 63 | Ghana | Policymakers | In-person survey | Validity |
| Reckers-Droog 2021 | Willingness-to-pay | 2023 | The Netherlands | National sample | Online | Validity |
| Ubel 1996 (a) | Choice experiment | 169 | Pennsylvania, US | Local sample of jurors | Paper questionnaire | Validity |
| Whitty 2014 | Discrete choice experiment and best-worst scaling | 930 | Queensland, Australia | Local sample | Online | Reliability |
| Lancsar 2011 | Discrete choice experiment | 587 | England | National sample of the general population | Face-to-face using Computer Assisted Personal Interview (CAPI) | Framing effects |
| Petrou 2013 | Person trade-off | 2500 | United Kingdom | National sample | Online survey | Reliability and framing effects |
| Aidem 2017 | Focus groups and semi-structured interviews | 27 | Norway | Hospital administrators, policymakers, practitioners, seniors, and university students | In-person | Validity |
| Whitty 2015 | Discrete choice experiment | 1994 | Queensland and South Australia | Local sample | Online survey | Face validity |
| Cookson 2018 | Questionnaire with pairwise choices | 60 | York, England | Local sample | Face-to-face | Framing and cognitive effects |
| Ahlert 2017 | Questionnaire | 166 | Germany | Local sample of university students | In-person | Validity |
| Ali 2017 | Choice experiment | 135 | York, England | Local sample | Online and face-to-face | Framing/cognitive effects |
| Li 2022 | Questionnaire | 1862 | United States | Physicians, compared with the general public, an 'elite' subsample of Americans, and a nationwide sample of medical students | Online survey | Validity |
| Ubel 2001 | Questionnaire | 615 | Philadelphia, USA | Local sample of randomly selected jurors | In-person survey | Framing effects |
| Ubel 1999 | Choice experiment | 479 | Philadelphia, US | Local sample of jurors | In-person survey | Framing effects |
| Abásolo 2013 | Choice experiment | 1211 | Spain | National sample | Face-to-face interview | Framing effects |
| Schoon 2022 | Integrated citizens jury and discrete choice experiment | 27 | Taiwan | Local sample | Face-to-face | Validity |
| McKie 2019 | Small-group discussion and interview | 66 | Victoria, Australia | Local sample | Semi-structured, face-to-face, small-group discussion | Framing effects |
| Comerford 2023 | Experiment with pairwise choices | 495 | United States and United Kingdom | National sample | Online survey | Reliability and validity |
| Baltussen 2006 | Discrete choice experiment | 30 | Ghana | Health policy makers | In-person survey | Validity |
| Ubel 1996 (b) | Choice experiment | 169 | Pennsylvania, US | Prospective jurors | Face-to-face survey | Framing effects |
| Green 2009 | Choice experiment | 261 | Southampton, England | Local sample | Face-to-face interview | Validity |
| van Exel 2015 | Q methodology | 294 | Ten countries: Hungary, Palestine, Poland, Denmark, Norway, France, Sweden, the UK, the Netherlands, Spain | General public, national samples | In-person and online | Reliability |

Table 2 presents the characteristics of validity, reliability, and framing effects as assessed in equity-efficiency trade-off studies.

**Table 2.**
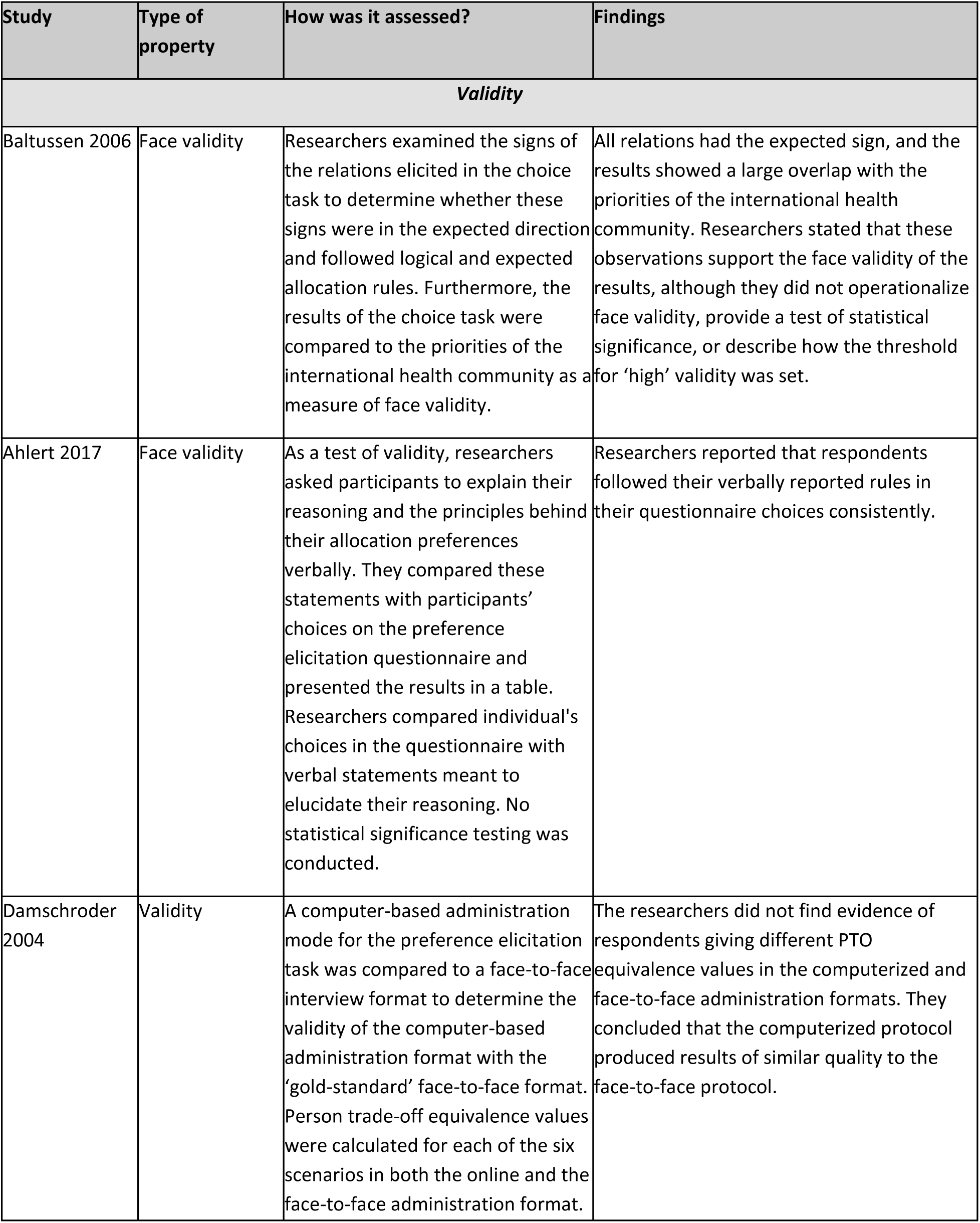
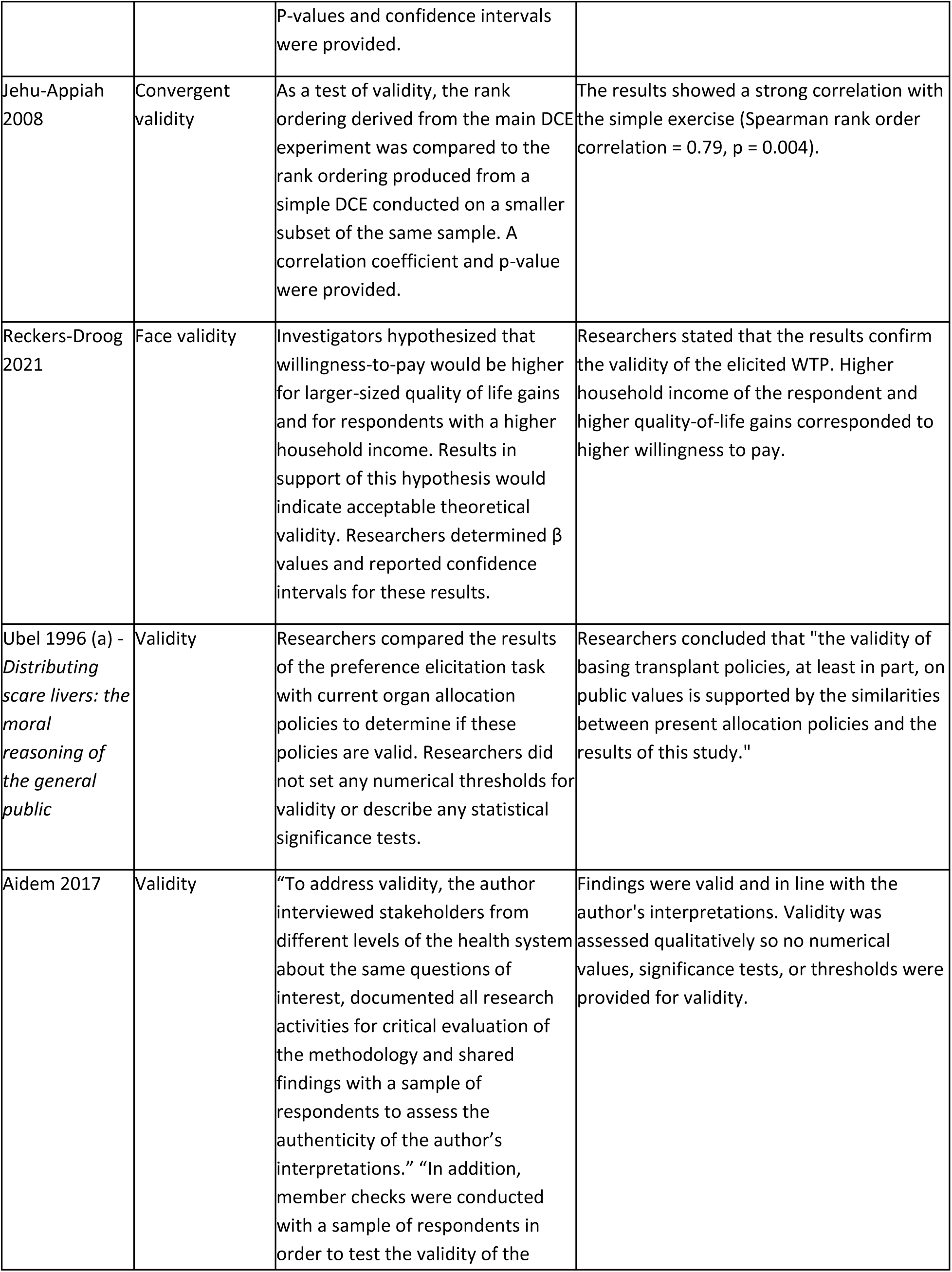
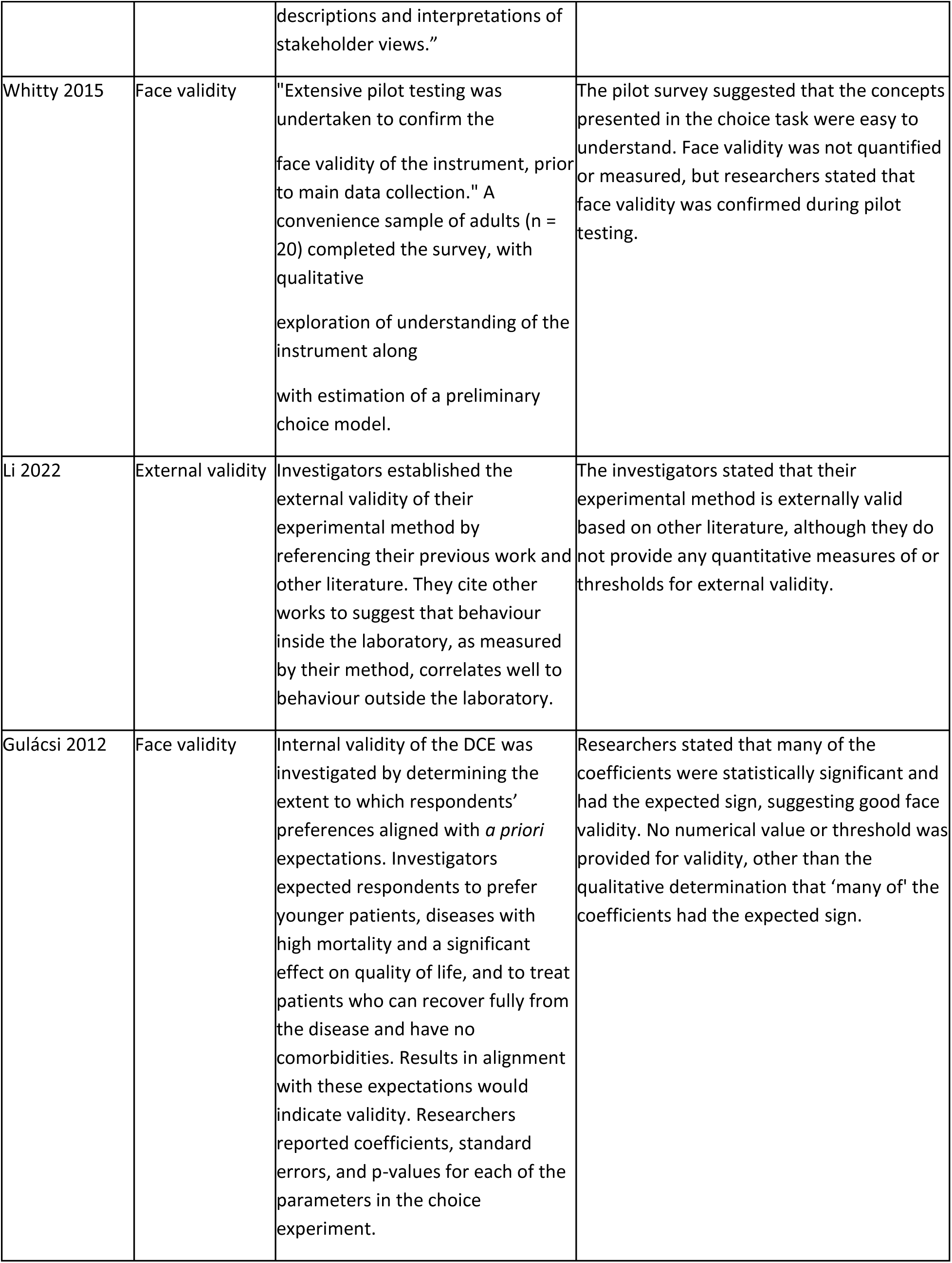
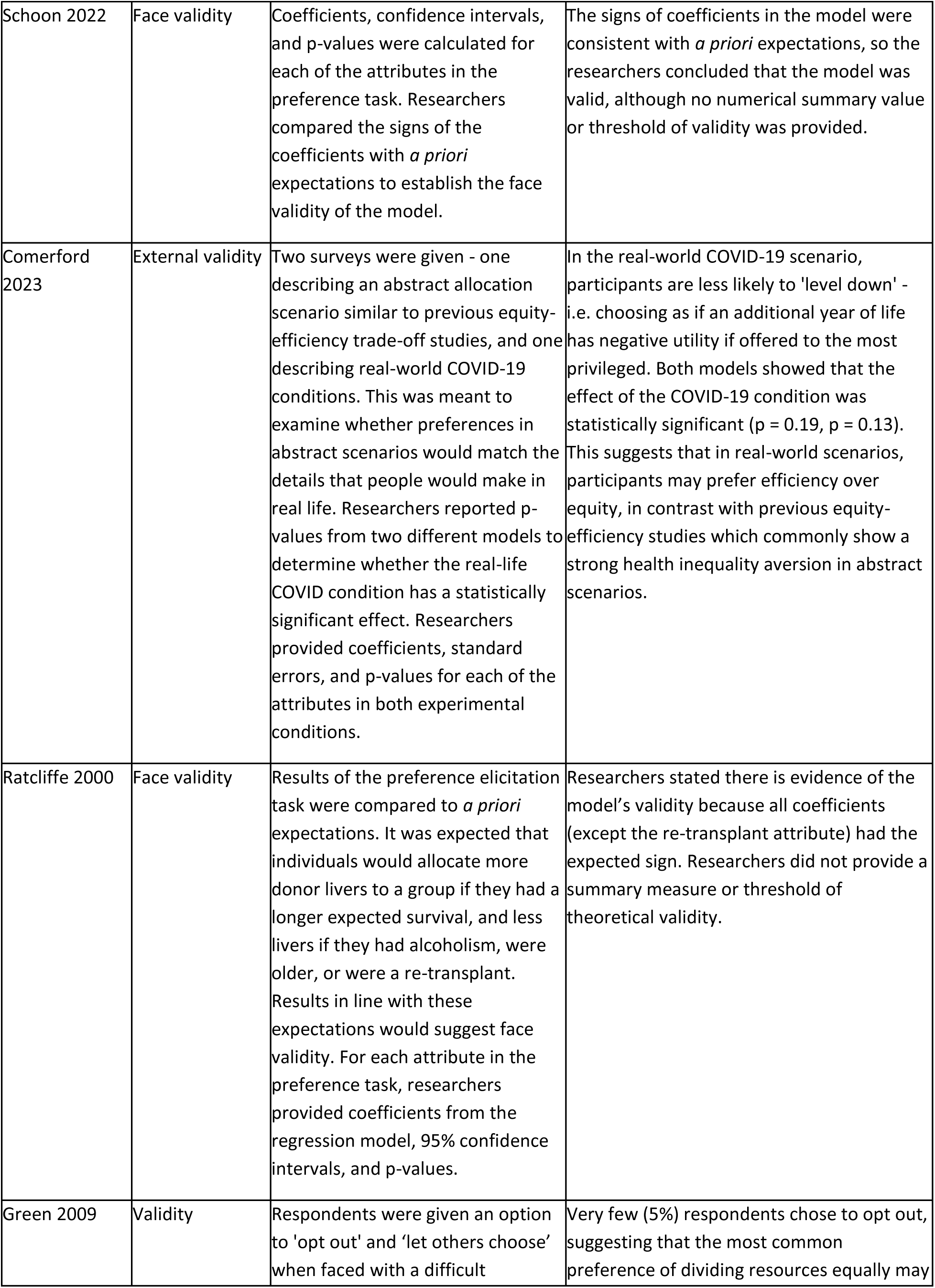
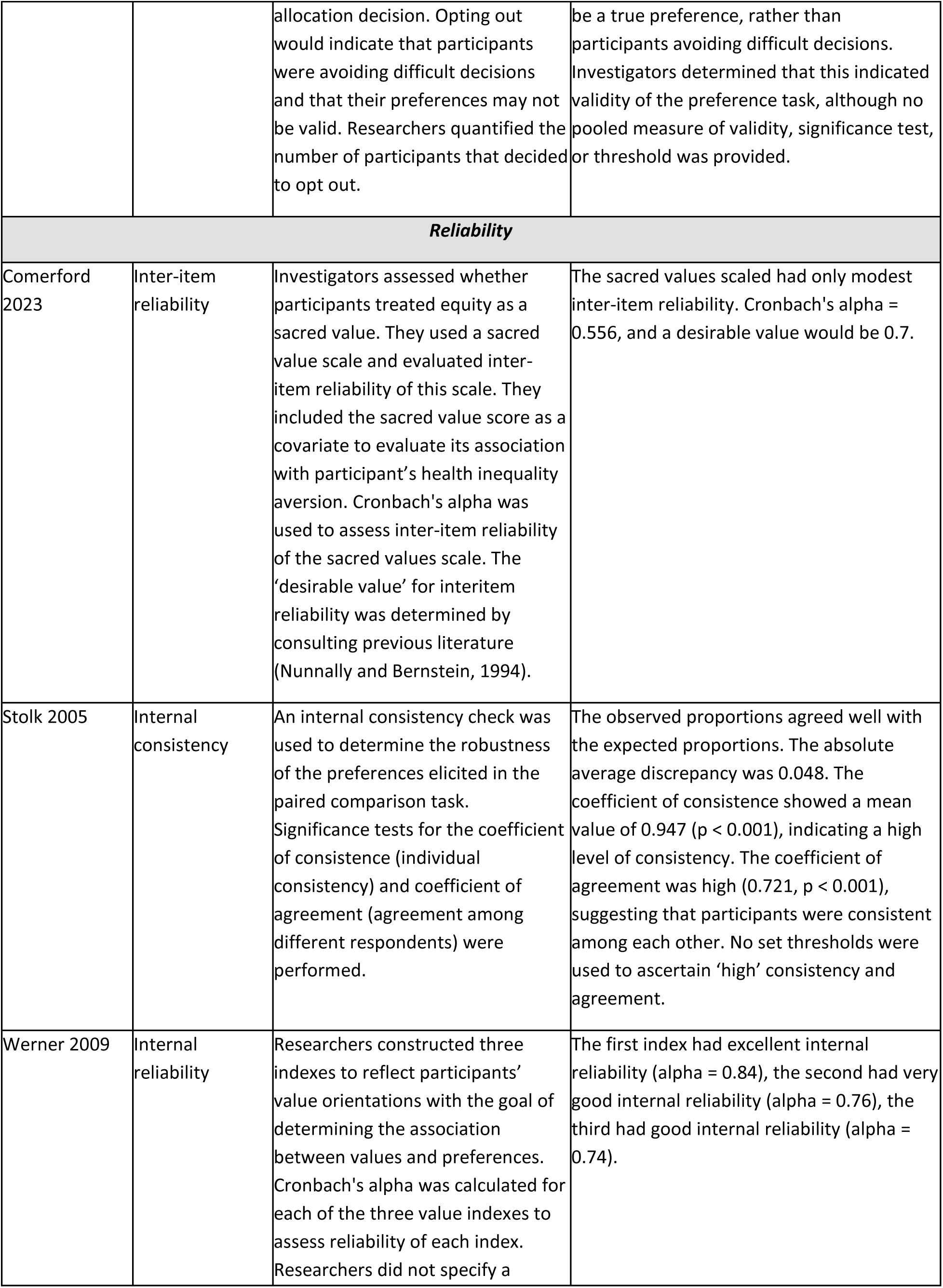
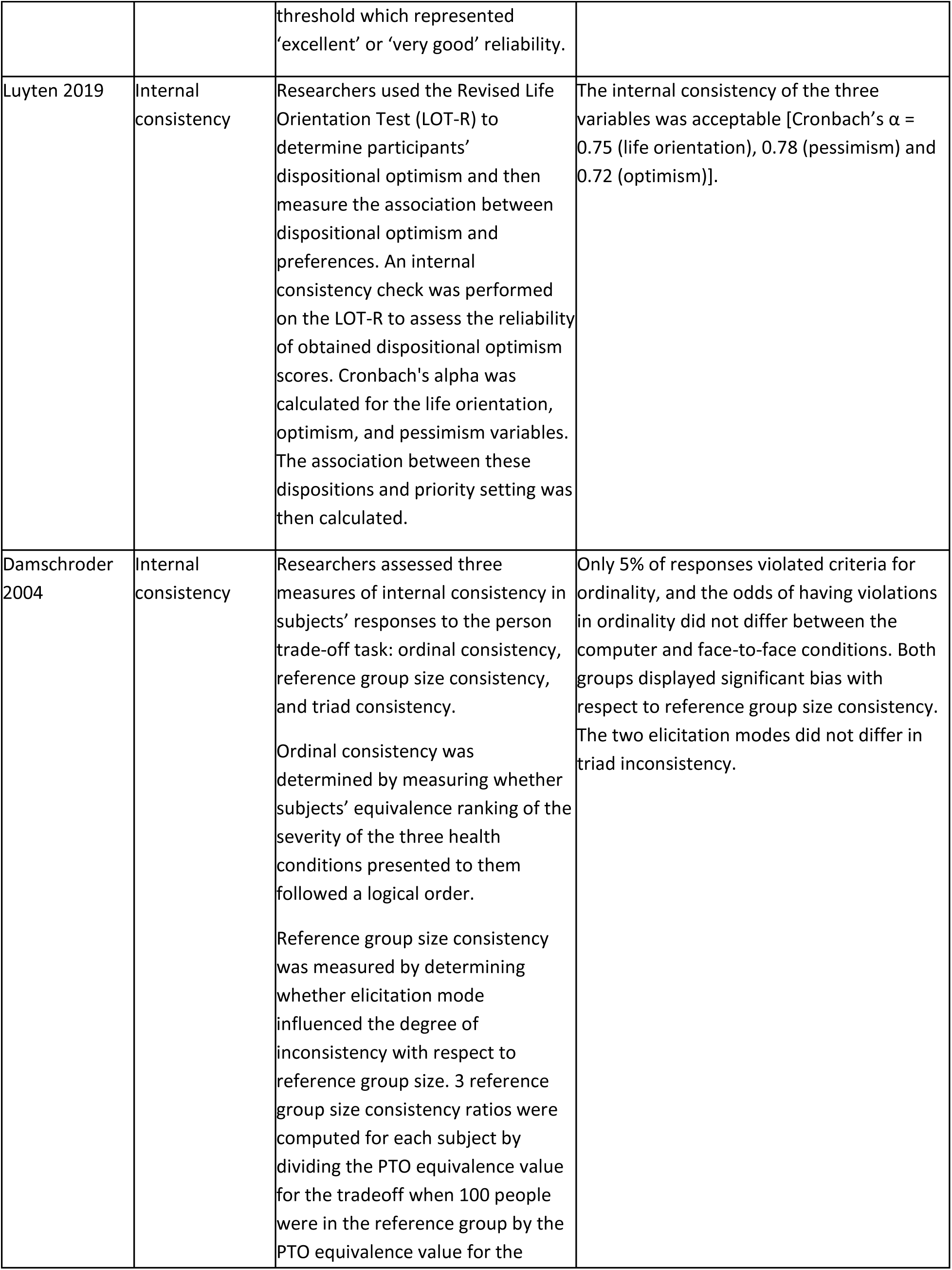
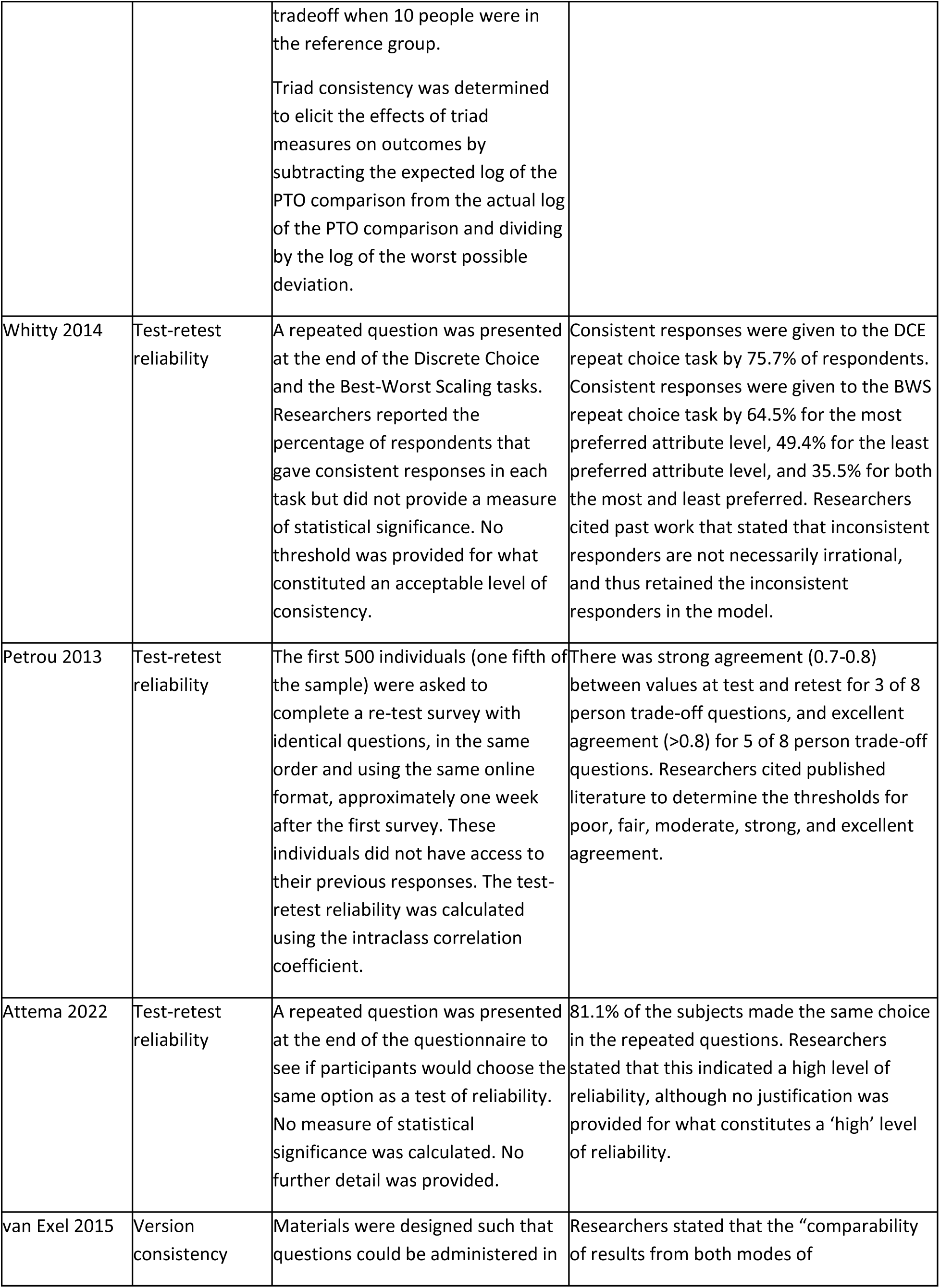
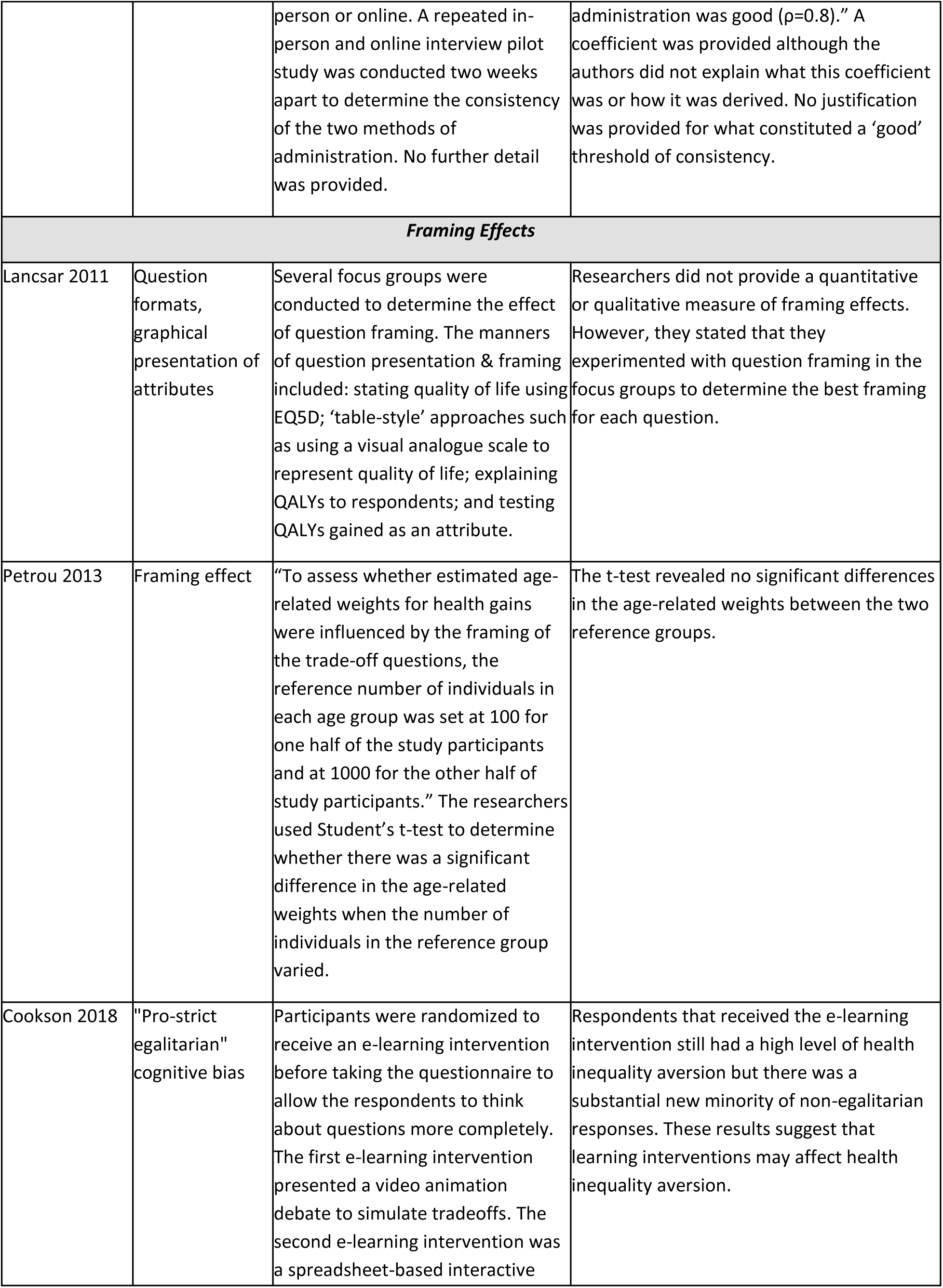
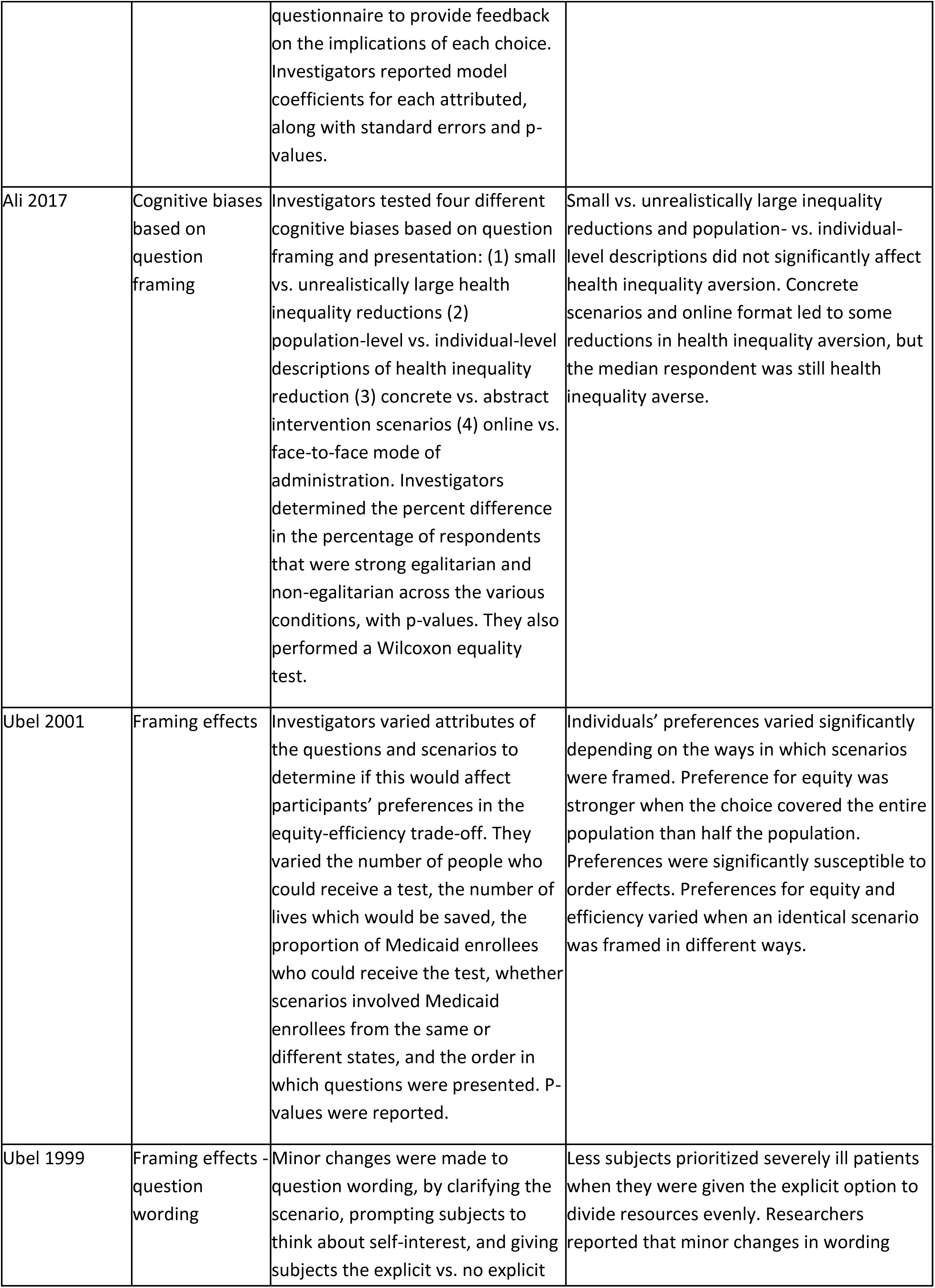
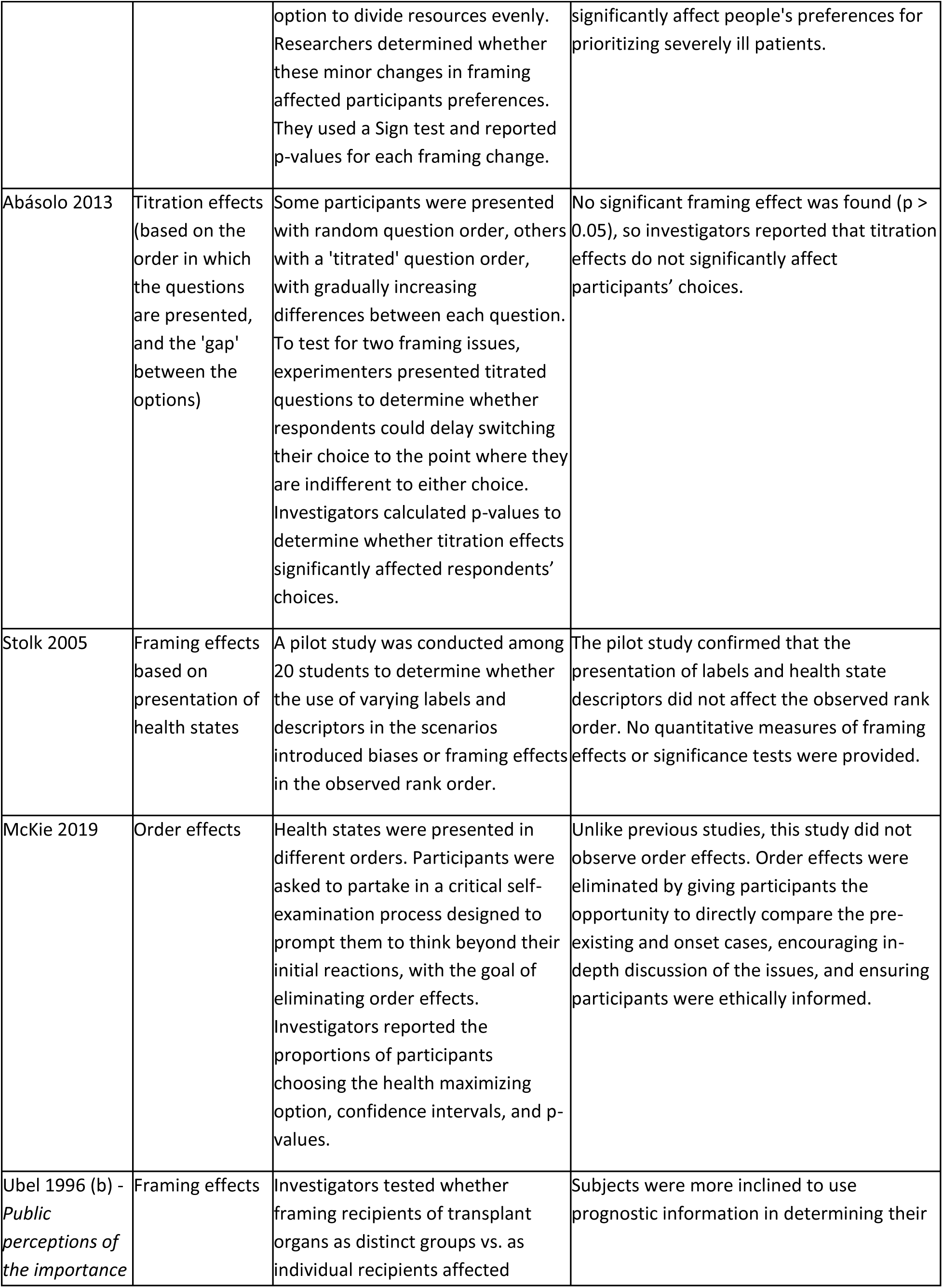
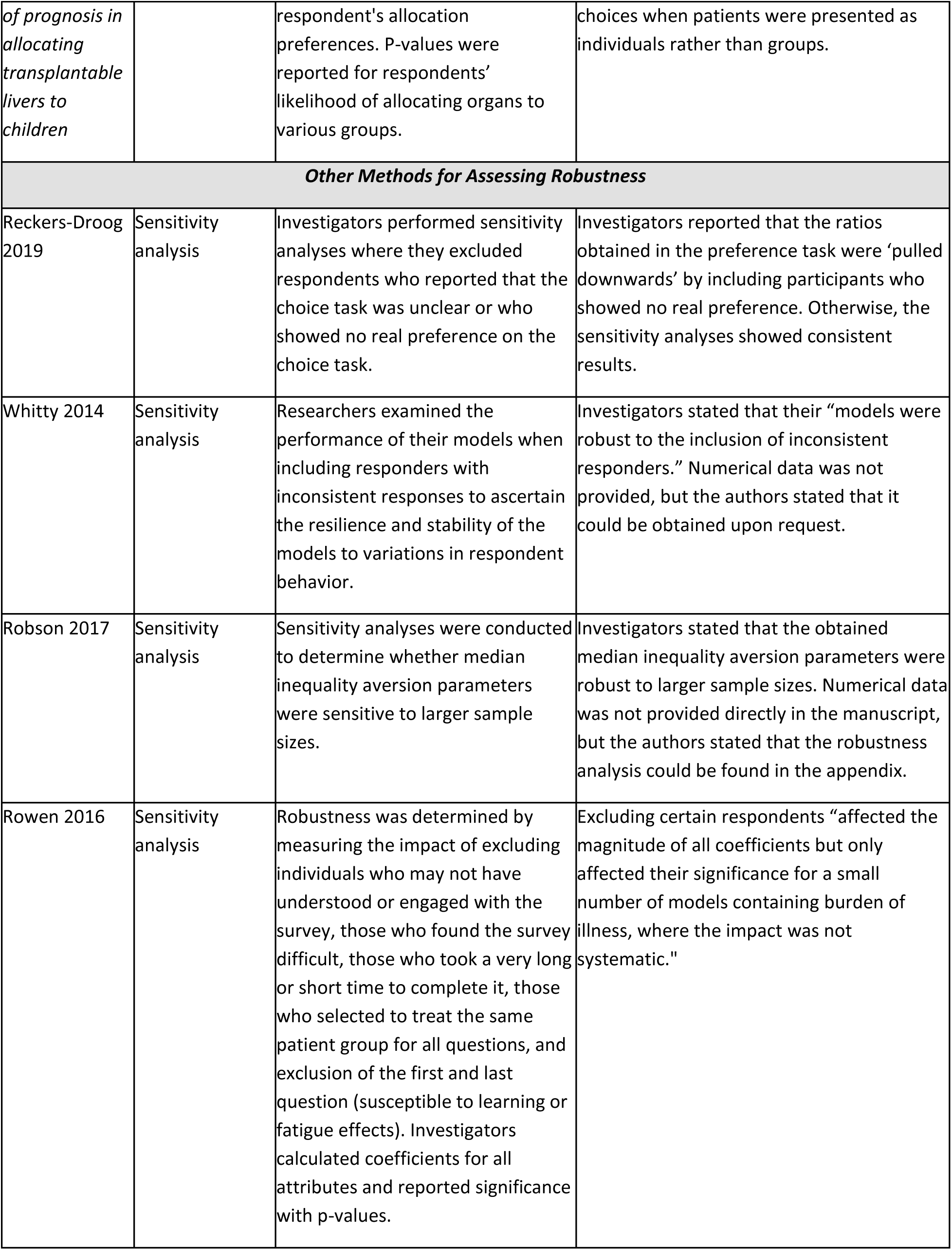
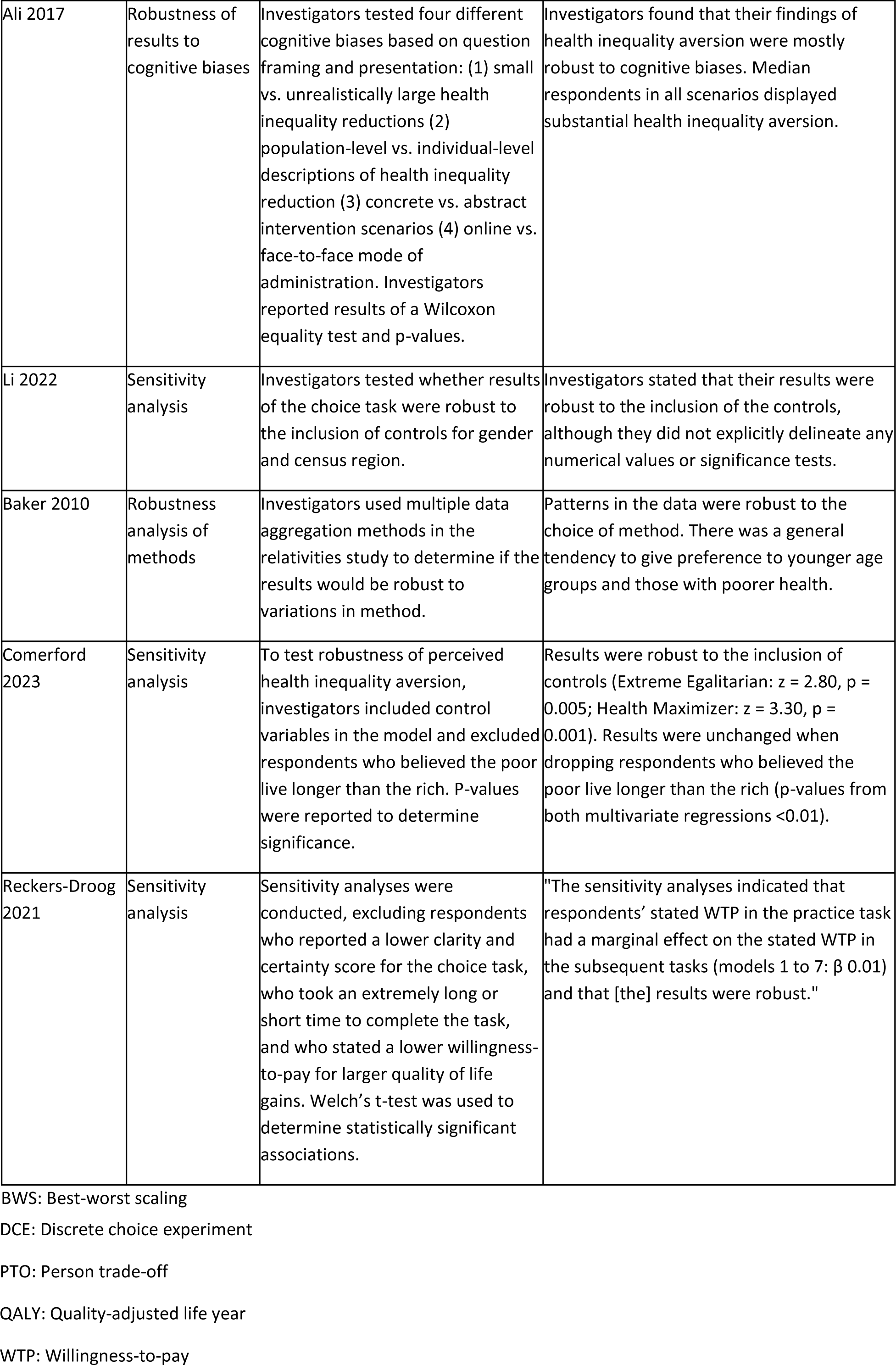
Characteristics of validity, reliability, and framing effects in equity-efficiency trade-off studies.

### Assessment of Validity

Fourteen studies assessed validity. Many studies did not explicitly specify the type of validity they assessed, although their assessment of validity could be aligned with definitions of face validity, convergent validity, or external validity. Several studies ascertained the face validity of their measurement instruments by comparing their results with *a priori* expectations or with previously published literature or policy,^50,53,55,60,61^ by conducting a pilot study to ascertain participant understanding,^49^ or by asking participants to verbally explain their reasoning.^37^ In all five studies that had *a priori* expectations for allocation preferences, researchers reported that results were in the hypothesized direction, suggesting good face validity. In the study that asked participants’ to explain their choices as a measure of face validity, investigators reported high validity because participants’ choices on the questionnaire were highly consistent with the verbal statements they made to explain their reasoning.^37^ Jehu-Appiah and colleagues assessed convergent validity by comparing the rank ordering from their main DCE exercise to the rank ordering produced from a simpler DCE exercise conducted on a subset of the sample. They found a strong correlation (Spearman rank order correlation = 0.79, *p* = 0.004) between the two DCE exercises.^39^ In a different study, researchers measured validity by giving participants the option to ‘opt out’ when faced with a difficult allocation decision.^46^ Only 5% of participants decided to opt out, suggesting that they were not avoiding difficult decisions and that their stated preferences reflected their true beliefs, indicating good validity.^46^ External validity was discussed in two equity-efficiency trade-off studies. Comerford and colleagues assessed external validity by comparing individuals’ preferences in an abstract scenario with their preferences in a real-world COVID-19 pandemic scenario. In the real-world scenario, participants were less likely to ‘level down’ and choose as if an additional year of life has negative utility if offered to the most privileged.^36^ This discrepancy in preferences between real-world and abstract resource allocation scenarios suggests that abstract scenarios may not be externally valid in eliciting individuals’ true preferences. Li and colleagues discussed external validity by referencing previous literature; they stated that their method, which measures behaviour in the laboratory, correlates well with behaviour outside of the laboratory.^44^ In the equity-efficiency studies that assessed validity, investigators did not explicitly reference monotonicity or compensatory preferences. Overall, most equity-efficiency studies reported acceptable validity of their measurement instruments.

### Assessment of Reliability

Of the nine studies that assessed reliability, five assessed reliability generally,^36,43,48,51,59^ three assessed test-retest reliability,^30,38,58^ and one assessed version consistency.^54^ In three of the studies assessing reliability, researchers constructed indices to determine participants’ value orientations, such as a dispositional optimism index, with the goal of including participants’ scores on the index as a covariate in the model of resource allocation preferences. To assess the reliability of these indices, researchers used Cronbach’s alpha, with two studies reporting good internal consistency of their indices^48,51^ and one study reporting modest internal consistency.^36^ To assess test-retest reliability, researchers provided repeated choice tasks^30^ or a repeated question at the end of the questionnaire.^58^ In another study, a subsample of individuals was asked to repeat the survey one week after the initial survey.^38^ Generally, researchers reported high test-retest reliability. For example, in one study, 81.1% of respondents made the same choices when provided with a repeated question,^58^ and in another study, there was strong or excellent agreement between the values provided at the initial test and at retest.^38^ To assess version consistency, van Exel and colleagues administered their survey both online and in-person two weeks apart and compared the results from both modes of administration.^54^ They stated that the comparability of results from both modes of administration was good.^54^ Level recoding was not explicitly mentioned in any of the equity-efficiency trade-off studies that discussed reliability.

### Assessment of Framing and Cognitive Effects

Ten studies assessed framing or cognitive effects. Half of these studies varied their question or attribute framing,^14,26,38,41,45^ and others assessed order effects by presenting questions in different orders.^47,56^ Two studies used a pilot study or focus groups to rule out framing and order effects prior to the main investigation.^40,43^ Five studies did not observe any significant framing or order effects.^14,38,43,47,56^ Conversely, three studies found that respondents’ choices were affected by question framing or order.^26,41,45^ In one study, investigators attempted to reduce pro-egalitarian cognitive bias by randomizing participants to receive an e-learning intervention designed to prompt participants to think about questions in a more complete manner prior to taking the questionnaire.^42^ Respondents that received the e-learning intervention still displayed high levels of inequality aversion, but there was a substantial new minority of non-egalitarian responses.^42^ In another study, Ali and colleagues tested four different cognitive biases based on question framing. They found that presenting small vs. unrealistically large inequality reductions in their hypothetical scenarios, and population-vs. individual-level descriptions did not affect health inequality aversion.^14^ The presentation of concrete scenarios instead of abstract scenarios, and an online administration format led to some reductions in health inequality aversion, but the median respondent was still inequality averse.^14^ In summary, while some studies reported that there were no major framing effects present in their study, others reported that framing effects significantly affected respondents’ choices, and others reported only a modest framing effect. As such, it is unclear to what extent framing effects influence the results of equity-efficiency trade-off studies.

### Other Methods of Assessing Robustness

In addition to validity, reliability, and framing effects, we identified other methods that equity-efficiency trade-off studies used to assess the robustness of their results. Several studies reported sensitivity analyses as a method of checking robustness.^16,29,61^ Three studies assessed robustness by repeating their analyses after excluding a certain group of participants. One study excluded participants who showed no real preferences in the choice task.^29^ Investigators in this study reported that their results were affected by excluding participants who showed no real preference.^29^ Another study repeated analyses after excluding participants who may not have understood or engaged with the survey, found the survey difficult, took a long or short time to complete the survey, or selected the same response for all questions.^31^ This study found that excluding certain participants affected the magnitude of all calculated coefficients, but this effect was only statistically significant for a small number of models and the impact was not systematic.^31^ A third study excluded participants who reported a lower clarity or certainty score for the choice task, took a long or short time to complete the task, or stated a lower willingness-to-pay for larger quality of life gains.^61^ Other equity-efficiency trade-off studies assessed robustness by including individuals with inconsistent responses^30^ and control participants.^36,44^ These studies found that results were robust to the inclusion of inconsistent responders and controls. One study assessed whether median inequality aversion parameters were sensitive to larger sample sizes and found that these parameters were indeed robust to sample size.^16^ In general, most studies reported that their results were robust to the inclusion or exclusion of certain participants, variations in methods, and differences in sample size.

The results of study quality assessment using the PREFS checklist can be found in Table 3.

**Table 3.** Study quality assessment of studies that assessed validity, reliability, or framing effects using the PREFS checklist.

| Study | Purpose (0-1) | Respondents (0-1) | Explanation (0-1) | Findings (0-1) | Significance (0-1) | Total (0-5) |
| --- | --- | --- | --- | --- | --- | --- |
| Werner 2009 | 1 | 0 | 1 | 1 | 1 | 4 |
| Luyten 2019 | 1 | 0 | 1 | 1 | 1 | 4 |
| Damschroder 2004 | 1 | 0 | 1 | 0 | 1 | 3 |
| Jehu-Appiah 2008 | 1 | 0 | 1 | 1 | 1 | 4 |
| Reckers-Droog 2021 | 1 | 0 | 1 | 0 | 1 | 3 |
| Ubel 1996(a) | 1 | 0 | 1 | 1 | 0 | 3 |
| Whitty 2014 | 1 | 0 | 1 | 1 | 1 | 4 |
| Lancsar 2011 | 1 | 0 | 1 | 0 | 1 | 3 |
| Petrou 2013 | 1 | 0 | 1 | 1 | 1 | 4 |
| Aidem 2017 | 1 | 0 | 1 | 1 | 0 | 3 |
| Whitty 2015 | 1 | 0 | 1 | 1 | 1 | 4 |
| Cookson 2018 | 1 | 0 | 1 | 1 | 1 | 4 |
| Ahlert 2017 | 1 | 0 | 1 | 0 | 0 | 2 |
| Ali 2017 | 1 | 0 | 1 | 1 | 1 | 4 |
| Li 2022 | 0 | 1 | 1 | 0 | 1 | 3 |
| Ubel 2001 | 1 | 0 | 1 | 0 | 1 | 3 |
| Gulácsi 2012 | 1 | 0 | 1 | 1 | 1 | 4 |
| Ubel 1999 | 1 | 0 | 1 | 1 | 1 | 4 |
| Abásolo 2013 | 1 | 0 | 1 | 0 | 1 | 3 |
| Schoon 2022 | 1 | 0 | 1 | 1 | 1 | 4 |
| Attema 2022 | 1 | 0 | 1 | 0 | 1 | 3 |
| Stolk 2005 | 1 | 0 | 1 | 1 | 1 | 4 |
| McKie 2019 | 1 | 0 | 1 | 1 | 1 | 4 |
| Comerford 2023 | 1 | 0 | 1 | 0 | 1 | 3 |
| Baltussen 2006 | 1 | 0 | 1 | 1 | 1 | 4 |
| Ubel 1996(b) | 1 | 0 | 1 | 1 | 1 | 4 |
| Ratcliffe 2000 | 1 | 0 | 1 | 0 | 1 | 3 |
| Green 2009 | 1 | 0 | 1 | 1 | 1 | 4 |
| van Exel 2015 | 1 | 0 | 1 | 0 | 1 | 3 |

## Discussion

This systematic review investigated whether and how equity-efficiency trade-off studies assess validity, reliability, or framing effects. Of the 115 included studies, only 29 (25.2%) did so. Less than 15% of the studies assessed validity and less than 10% of the studies assessed reliability or framing effects. In the studies that assessed them, investigators used a wide array of quantitative and qualitative assessment methods. Face validity was commonly assessed by comparing results to hypothesized expectations. Convergent validity was assessed in one study where researchers provided a simple version of the exercise to a subsample of participants and compared the results of the simple exercise to those of the main choice experiment. External validity was assessed in one study by comparing abstract and real-world scenarios, and in another study by referencing previously published literature. Level recoding and compensatory preferences were not explicitly referenced in any of the studies that discussed validity or reliability. Researchers assessed test-retest reliability and version consistency by offering a repeated choice task or by offering a different version of the questionnaire. Reliability was also assessed by determining the internal consistency of indices used to ascertain participants’ value orientations. Generally, equity-efficiency trade-off studies reported good reliability and validity, with fewer studies reporting low validity and reliability. Multiple studies reported no framing and cognitive effects, while others reported that their results were significantly affected by framing and cognitive effects. Studies that used other methods for assessing the robustness of their results, such as sensitivity analyses, reported that their results were robust to variations in method or exclusion of certain respondents.

To our knowledge, this is the first systematic review to discuss the validity, reliability, and framing effects of equity-efficiency trade-off studies. Several review papers have assessed the validity of DCEs and other preference elicitation techniques. Merlo and colleagues conducted a validity assessment of DCEs of primary healthcare professionals, with specific assessment of internal and external validity.^17^ A meta-analysis by Quaife and colleagues assessed external validity of health-related DCEs to determine how well DCEs predict health choices in real life.^144^ Other reviews have investigated validity and concordance of DCEs and best-worst scaling methods in health,^34^ participant understanding in health-related DCEs,^145^ and external validity of DCEs in health economics.^146^ A systematic review by Ryan and colleagues described preference elicitation techniques in the health literature, including their validity, reliability, and generalizability.^147^ Although these reviews discuss the validity and reliability of preference elicitation techniques, they review health-related DCEs more broadly, without a specific focus on equity as a dimension of choice.

It is unsurprising that few equity-efficiency trade-off studies report validity, reliability, and framing effects. A previous systematic review of self-report research utilization measures in healthcare found that only 33% of studies assessed reliability.^148^ The authors of this review noted that there is significant underdevelopment in psychometric assessment.^148^ Although equity-efficiency trade-off studies are not psychometric assessments, these conclusions from the assessment of psychometric literature suggest that validity and reliability may be even more underreported in preference elicitation choice experiments, where validity and reliability are not traditionally systematically assessed. The findings of our systematic review support this conclusion. It appears that validity and reliability are underreported across health research disciplines.

Previous studies have reported mixed findings about the validity and reliability of preference elicitation techniques. A systematic review and meta-analysis of external validity of DCEs found that DCEs have moderate, but not exceptional, accuracy in predicting health choices.^144^ Conversely, a preference elicitation study for health states that used visual analogue scale, time trade-off, and standard gamble methods found that these methods had poor test-retest reliability and construct validity.^149^ Another study showed that DCEs are able to accurately predict health choices, mimicking real-world decisions, if scale and preference heterogeneity are considered.^150^ Given these differing findings in the health economics literature, it is unsurprising that we found varying reports of validity and reliability in equity-efficiency trade-off studies, with some studies reporting high or moderate validity and reliability and other studies reporting poor validity and reliability. These conflicting findings may be due to differing preference elicitation methods, study samples, or other context-specific factors.

Only a small proportion of equity-efficiency trade-off studies assessed or discussed the validity, reliability and framing effects of their measurement instruments. This finding is concerning, given that the purpose of equity-efficiency trade-off studies is to inform high-impact health policy and health resource allocation decisions. Researchers in other fields have noted that reporting validity and reliability is necessary to evaluate the usefulness of measurement instruments and meaningfully apply results to clinical practice and policy.^151^ Establishing the validity and reliability of stated preference methods is important if the results of studies are to be used to inform health policy.^146^ Assessment of validity and reliability lends credibility to the method used.^152^ The results of these studies must be interpreted critically and cautiously if there is no such assessment.

Researchers should assess validity and reliability every time they are conducting a new equity-efficiency trade-off study, even if they are using a preference elicitation technique for which validity and reliability have previously been established. Validity and reliability are not fixed and depend on study context, study population, and other factors.^23^ Although several studies have assessed the validity of common preference elicitation techniques such as discrete choice experiments,^17,144^ these assessments are not specific to the equity-efficiency trade-off. When conducting an equity-efficiency trade-off study, it is not sufficient to cite previous studies and suggest that all discrete choice experiments are valid. The validity and reliability of preference elicitation techniques will vary greatly depending on the context in which they are applied.

A limitation of this systematic review is that the assessment methods of included studies were heterogeneous. Studies assessed validity and reliability using different methods and reported a wide range of qualitative and quantitative measures. Moreover, studies varied widely in their preference elicitation techniques, with methods ranging from discrete choice experiments to person trade-off to integrated citizens juries and others. This heterogeneity of study methods precluded a meta-analysis or a direct comparison of validity and reliability across equity-efficiency trade-off studies. This review serves as a descriptive paper to outline the current state of validity and reliability assessment and reporting in the equity-efficiency literature.

Another limitation of this systematic review is that study reviewers were not blinded to study outcomes. As such, a selection bias is possible if reviewers were more inclined to include a study if it mentioned validity or reliability in the title or abstract. Although such a selection bias is possible, it was likely mitigated due to duplicate study screening by two independent reviewers and resolution of conflicts by a third reviewer.

Currently, researchers use a wide array of preference elicitation techniques in their equity-efficiency trade-off studies, such as discrete choice experiments, person trade-off, best-worst scaling, and many others. Future studies should compare the validity and reliability of these various preference elicitation methods directly to ascertain whether certain preference elicitation methods are more valid and reliable than others in deriving preferences for equity and efficiency.

Another promising avenue for future research is determining a ‘threshold’ for validity and reliability in equity-efficiency trade-off studies, if such a threshold exists. Researchers and policy makers should consider how high validity and reliability of an equity-efficiency trade-off study need to be for this study to be considered acceptable for use in health policy decision making.

Researchers may be unaware of the need to assess the validity and reliability of their methods. As such, future work should focus on developing guidelines to support researchers in the transparent assessment and reporting of validity and reliability in their studies. Such guidelines may outline minimum standards for the reporting of validity and reliability in preference elicitation studies.

## Conclusion

This systematic review described validity, reliability, and framing effects in equity-efficiency trade-off studies. Most equity-efficiency trade-off studies did not assess validity, reliability, or framing effects. The studies which assessed them presented a wide range of qualitative and quantitative metrics, with most studies reporting good validity and reliability and fewer reporting low validity and reliability. The results of equity-efficiency trade-off studies should be interpreted with caution if there is no assessment of important measures such as validity and reliability. Equity-efficiency trade-off studies should only be used to inform high-impact health policy decisions if authors demonstrate acceptable validity and reliability.

## Data Availability

All data produced in the present study are available upon reasonable request to the authors.

