## Appendix A for "Validity, Reliability, and Framing Effects in Equity-Efficiency Trade-Off Studies: A Systematic Review"

**Appendix A:** Full Database Search Strategies

***MEDLINE***

1 health.tw,kf.
2 (healthcare or health-care or 'health care').tw,kf.
3 *Health Priorities/
4 *Health Care Rationing/
5 *"Value of Life"/
6 *Quality-Adjusted Life Years/
7 (QALY or "quality-adjusted life year$").tw,kf.
8 *Resource Allocation/
9 1 or 2 or 3 or 4 or 5 or 6 or 7 or 8
10 ('choice behaviour' or 'choice behavior' or 'choice experiment*' or 'value elicit*' or DCE or 'person trade-off$' or 'person tradeoff$' or PTO or 'conjoint analysis' or 'preference* elicit*' or 'trade-off$' or 'tradeoff$').tw,kf.
11 ('stated preference*' or 'public preference*' or 'community preference*' or 'societal preference*' or 'priority setting').tw,kf.
12 ('social value$' or 'societal value$' or (distribut* adj2 preference*) or 'social choice' or 'relative value$' or 'community value$').tw,kf.
13 *Choice Behavior/
14 10 or 11 or 12 or 13
15 (inequalit* or equit* or inequit*).tw,kf.
16 ((distribution* adj weight*) or "equity weight*" or (QALY adj2 weight*) or (equity adj2 preference*) or "lifetime health" or (QALY and "relative value")).tw,kf.
17 ((health adj maximi*) or "health benefit maximi*").tw,kf.
18 ("outcome egalitaria*" or "gain egalitaria*" or prioritaria* or sufficientaria* or Rawlsian).tw,kf.
19 ('Social Welfare Function*' or SWF).tw,kf.
20 'inequality aversion'.tw,kf.
21 ("fair innings" or "egalitarian ageism" or "age-related weights" or "age-weighting preferences").tw,kf.
22 ('absolute shortfall' or 'proportional shortfall').tw,kf.
23 15 or 16 or 17 or 18 or 19 or 20 or 21 or 22
24 9 and 14 and 23

***EMBASE***

1 health.tw,kf.
2 (healthcare or health-care or 'health care').tw,kf.
3 "health priorities".tw,kf.
4 "health care rationing".tw,kf.
5 "value of life".tw,kf.
6 "quality-adjusted life years".tw,kf. or *quality adjusted life year/
7 "resource allocation".tw,kf. or *resource allocation/
8 1 or 2 or 3 or 4 or 5 or 6 or 7
9 ('choice behaviour' or 'choice behavior' or 'choice experiment*' or 'value elicit*' or DCE or 'person trade-off$' or 'person tradeoff$' or PTO or 'conjoint analysis' or 'preference* elicit*' or 'trade-off$' or 'tradeoff$').tw,kf.
10 ('stated preference*' or 'public preference*' or 'community preference*' or 'societal preference*' or 'priority setting').tw,kf.
11 ('social value$' or 'societal value$' or (distribut* adj2 preference*) or 'social choice' or 'relative value$' or 'community value$').tw,kf.
12 9 or 10 or 11
13 (inequalit* or equit* or inequit*).tw,kf.
14 ((distribution* adj weight*) or "equity weight*" or (QALY adj2 weight*) or (equity adj2 preference*) or "lifetime health" or (QALY and "relative value")).tw,kf.
15 ((health adj maximi*) or "health benefit maximi*").tw,kf.
16 ("outcome egalitaria*" or "gain egalitaria*" or prioritaria* or sufficientaria* or Rawlsian).tw,kf.
17 ('Social Welfare Function*' or SWF).tw,kf.
18 'inequality aversion'.tw,kf.
19 ("fair innings" or "egalitarian ageism" or "age-related weights" or "age-weighting preferences").tw,kf.
20 ('absolute shortfall' or 'proportional shortfall').tw,kf.
21 13 or 14 or 15 or 16 or 17 or 18 or 19 or 20
22 8 and 12 and 21

***Web of Science***

"health priorit*" or "health$care rationing" or "value of life" or "quality-adjusted life year$" or "resource allocation" or "health" or "health$care" or "health benefit*"

AND

"choice behavior" or "choice behaviour" or "choice experiment*" or "value elicit*" or DCE or "person trade-off$" or "person tradeoff$" or PTO or "conjoint analysis" or "preference* elicit*" or "trade-off$" or "tradeoff$" or "stated preference*" or "public preference*" or "community preference*" or "societal preference*" or "priority setting" or "social value$" or "societal value$" or (distribut* adj2 preference*) or "social choice" or "relative value$" or "community value$" or "rationing scenario" or "attitude to health"

AND

"equalit*" or "inequalit*" or "equit*" or "inequit*" or (distribution* adj weight*) or "equity weight*" or (QALY adj2 weight*) or (equity adj2 preference*) or (QALY and relative value) or (health adj maximi*) or "health benefit maximi*" or egalitaria* or prioritaria* or sufficientaria* or "health inequality aversion" or "social welfare function" or SWF or "fair innings" or "ageism" or "age-related weights" or "age-weighting preferences" or "absolute shortfall" or "proportional shortfall" or (preference adj3 "severely illl") or "health prospect*" or "health status disparities" or "severity of illness index" or "socioeconomic factors"
